# In praise of fossil fuel subsidies (for cooking) ^‡^

**DOI:** 10.1101/2023.10.26.23297550

**Authors:** Carlos F. Gould, Rob Bailis, Kalpana Balakrishnan, Marshall Burke, Sebastián Espinoza, Sumi Mehta, Samuel B. Schlesinger, José R. Suarez-Lopez, Ajay Pillarisetti

## Abstract

Households that burn biomass in inefficient open fires – a practice that results in $1.6 trillion in global damages from health impacts and climate-altering emissions yearly – are often unable to access cleaner alternatives, like gas, which is widely available but unaffordable, or electricity, which is unattainable for many due to insufficient supply and reliability of electricity services. Governments are often reluctant to make gas affordable. We argue that condemnation of all fossil fuel subsidies is short-sighted and does not adequately consider subsidizing gas for cooking as a potential strategy to improve public health and reduce greenhouse gas emissions.

---

Fossil fuel subsidies are broadly condemned as economically and environmentally detrimental. We argue that, for the case of subsidizing fossil fuels for cooking, this condemnation is myopic and can deepen health and energy inequities. Subsidizing gas for cooking can help more than 400 million poor, marginalized, and vulnerable households avoid the large health and climate costs associated with traditional biomass cooking.

Many – including policy actors, economists, and environmentalists – contend that subsidies, especially for fossil fuels, are an inefficient allocation of resources that generate large fiscal burdens, are disproportionately captured by the wealthy, and that crowd out better policy and financial alternatives ^1,2^. We agree. Fossil fuels harm the climate. They account for 75% of global greenhouse gas emissions ^3^. Fossil fuels also harm human health. They are responsible for millions of premature deaths yearly from ambient air pollution ^4^.

Of course, fossil fuels have also powered modern life for well over a century. However, recent scale up of reliable, low-cost, and clean renewable energy indicates that these energy sources can facilitate continued economic growth and reduce the harms that fossil fuels cause to planetary and human health. The distribution of these benefits is, unsurprisingly, unequal; for much of the world, this clean energy future is a long way off. Today, biomass (firewood, dung, crop residues, charcoal) combustion in traditional stoves for cooking and heating emits pollution responsible for 4% of all premature deaths and 2% of global carbon dioxide equivalent (CO_2_e) emissions ^5,6^. Under current policy commitments, 30% of the global population will lack access to modern cooking fuels (gas and electricity) by 2030 ^7^. Waiting for existing policies or the free market to close these gaps forces marginalized populations to continue inefficient, polluting cooking practices. ^5,6^

Arguably, the most viable solution available in the near-term for reducing the harms of inefficient biomass combustion is liquified petroleum gas (LPG), a blend of propane and butane stored in stable, transportable cylinders. As compared to using biomass, those that cook with gas experience much lower air pollution exposures ^8,9^ and produce fewer greenhouse gas emissions when accounting for unsustainable wood harvesting ^10^. However, biomass-reliant households are often too poor to afford near-exclusive LPG use ^11^, which is required to substantially reduce air pollution exposures and improve health.

Gas subsidies can address multiple market failures that limit LPG adoption and use. Subsidies can alleviate financial constraints faced by biomass-dependent households resulting from low incomes or restricted access to credit. LPG subsidies also can reduce the myriad external costs of biomass combustion, including higher household and ambient air pollution and greenhouse gas emissions, adverse health outcomes, and landscape degradation. Information campaigns to promote the benefits of cleaner cooking, which could avoid or reduce subsidy deployment, have not been successful in increasing gas consumption. It is increasingly clear that costs are the primary barrier to widespread and near-exclusive LPG use ^11–13^. Still, the prospect of making cooking gas affordable through sustained subsidy mechanisms, even as a transitional fuel, is not widely considered a viable policy choice.

We use three country case studies in Ecuador, India, and Kenya to quantify the health and climate benefits accrued by reduced reliance on cooking with biomass fuels in the presence of affordable, subsidized LPG.

## Development and impact of long-standing gas subsidies in Ecuador

In the 1970s, Ecuador’s petroleum boom spurred government spending on welfare-enhancing programs to generate political support, including a universal subsidy for residential LPG that began in 1979. Per capita LPG consumption grew by an average of 31% annually in the 1970s, by 12% in the 1980s, and by 4% from 1990–2010. From 1979–2019, the LPG subsidy cost the government $13.3 billion (0.8% of GDP and 2-5% of government spending) (Table 1).

**Table 1.**
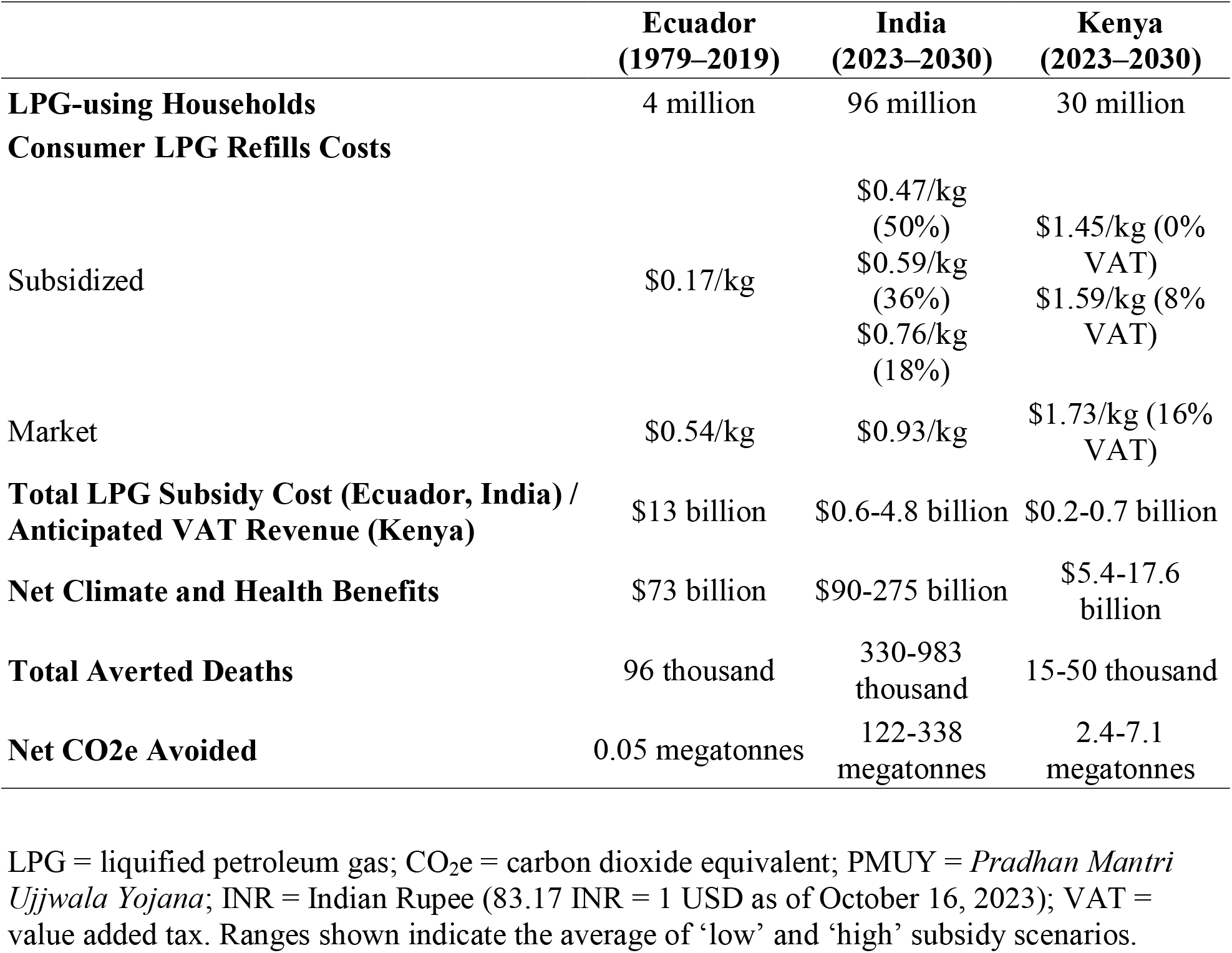
Health and climate impacts of LPG subsidies in Ecuador, India, and Kenya.

To model averted deaths from increased LPG adoption for cooking since 1979, we combine nationwide mortality rates, population counts, the fraction of households using a clean-burning fuel for cooking, personal fine particulate matter (PM_2.5_) exposures for cooking with firewood versus gas (Fig 1a), and existing PM_2.5_ exposure-mortality relationships ^14^ (Methods). We compare observed LPG scale-up to a counterfactual scenario where Ecuador’s transition is slowed by 20 years, mirroring adoption in neighboring Peru.

**Figure 1.**
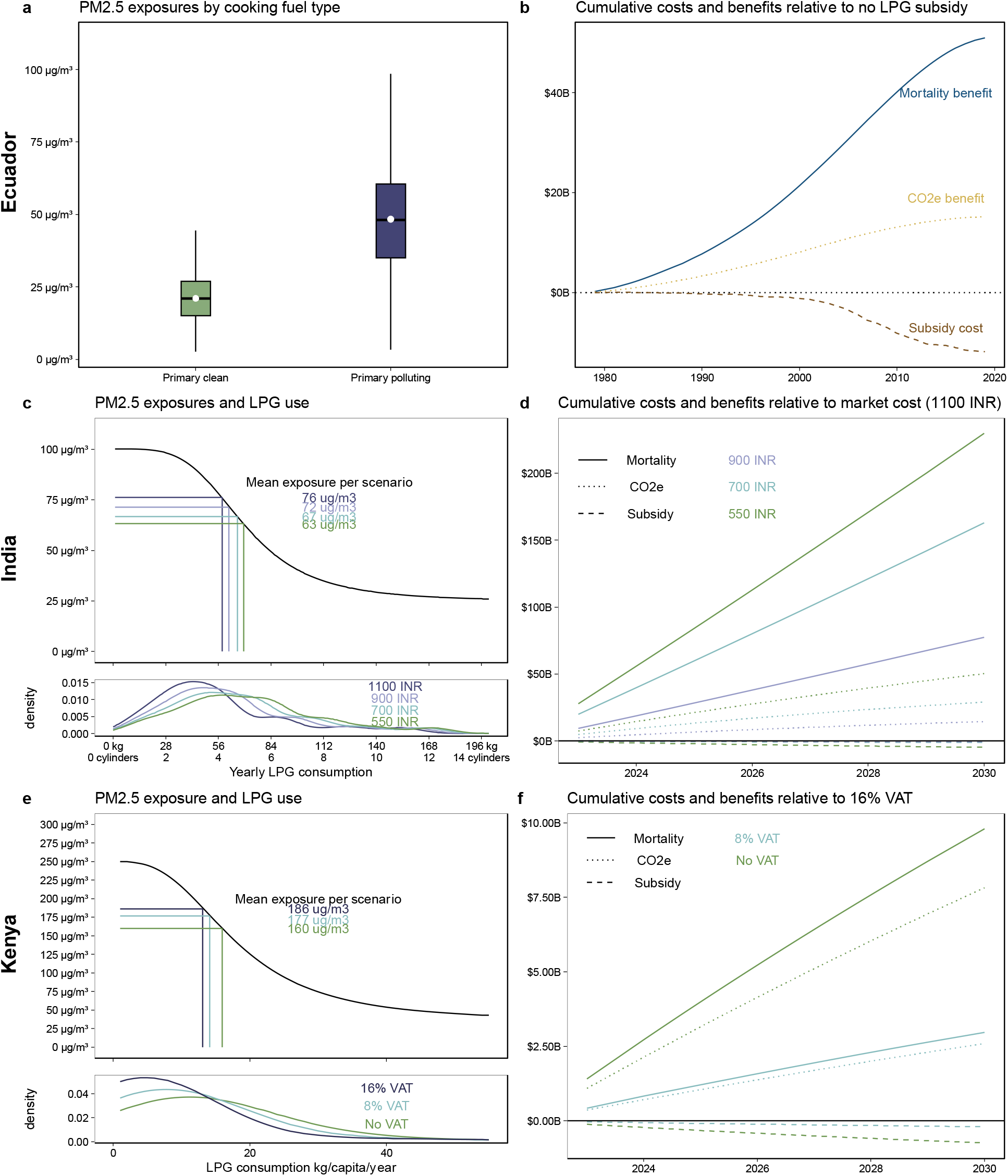
Benefits from LPG subsidies outweigh associated costs across three case studies in Ecuador, India, and Kenya. Panels **a and b** describe the data and results for Ecuador. Panel **a** shows the distribution of 1,000 average annual PM2.5 exposure estimates for those living in households that rely on clean burning fuels (namely gas) as compared to those that rely on biomass primarily. Panel **b** indicates calculated cumulative mortality benefits, CO2e-related benefits, and costs related to subsidizing gas in USD relative to the counterfactual scenario where there was no LPG subsidy. Panels **c and d** describe the data and results for India. Panel **c top panel** shows our modeled relationship between yearly LPG consumption per household and average annual PM2.5 exposure estimates for those living in those households across 1000 bootstrapped runs, shading indicates 2.5^th^-97.5^th^ lines at each 1 kg LPG increment across 1000 draws; annotations indicate the average exposure across the 1000 bootstrapped runs of each scenario. Panel c **bottom panel** indicates the density of households LPG consumption across each price scenario. Panel **d** shows calculated cumulative mortality benefits, CO2e-related benefits, and costs related to subsidizing gas in USD relative to the counterfactual scenario of no subsidies (i.e., one 14.2 kg LPG cylinder refill costs 1100 INR). Panels **e and f** describe the data and results for Kenya and mirror those of India where the counterfactual is a 16% VAT.

We calculate that 98,000 premature deaths were averted between 1979–2019 because of subsidy-induced accelerated LPG uptake. This estimate agrees with similar modeling from the Global Burden of Disease and with regression evidence that relates yearly mortality rates with cooking fuel use from 1990–2019 at the canton level (see SM). To quantify program impacts on CO_2_e, we apply standard estimates of energy demand and fuel emissions (Methods); we estimate that actual LPG scale-up avoided 52 kilotonnes CO_2_e from 1979–2019, i.e., 22% fewer cooking-related emissions in the country. Monetizing mortality and climate changes indicates that benefits from the LPG subsidy outweigh costs four to one (Table 1, Fig 1b).

## Benefits from maintaining the world’s largest LPG subsidy in India

More than 500 million Indians live in a home that recently acquired an LPG stove via the large-scale government program Pradhan Mantri Ujjwala Yojana (PMUY); nevertheless, an estimated two-fifths of Indian households continue to cook primarily with biomass. While PMUY has been rightly heralded as a success, access alone is not enough: sustained, near-exclusive clean cooking fuel use is necessary to maximize health and climate benefits. For LPG to be used regularly, it must be affordable. Historically, PMUY beneficiaries purchased cylinder refills at market rates. Subsidies, which can defray up to 45% of the market cost, are subsequently deposited into customers’ bank accounts. However, the Government of India almost entirely cut the 2023-2024 budget allocated to subsidizing LPG ^15^, dimming hopes for a more complete transition to clean fuels; as of July 2023, a fraction of subsidies for LPG consumers were restored.

To estimate the benefits of different levels of the LPG subsidy for PMUY beneficiaries, we consider three scenarios for the cost to consumers of an LPG cylinder refill – the 2019 subsidized cost (550 INR), the current subsidized cost (700 INR), and a more modest cost of 900 INR – each of which we compare to current market cylinder refill costs (1100 INR). We estimate kilograms of LPG consumed per year per household from nationally representative survey data collected in 2019. We map consumption to personal PM_2.5_ exposures, drawing on a recent clinical trial of a free LPG stove and fuel intervention where PM_2.5_ exposures were extensively measured ^8^ (Fig 1c). Using LPG price elasticities recovered from a randomized subsidy experiment in rural India among PMUY beneficiaries ^16^, we estimate that, with the LPG subsidy, LPG consumption increases and personal PM_2.5_ exposures decrease (Fig 1c). Using existing exposure-mortality risk curves ^14^ and population data from 2023-2030, we translate estimated increased PM_2.5_ exposures to changes in relative risks and predicted yearly crude mortality rates.

With even the smallest subsidy, we estimate an average of 330,000 premature deaths averted by 2030; averted deaths are three times larger when refills are subsidized down to 550 INR. For climate impacts, we predict that LPG subsidies would avoid 120-340 megatonnes CO_2_e. Relative to a price of 1,100 INR per cylinder refill, after applying a social discount rate of 9%, averted mortality benefits total $77-230 billion and avoided CO_2_e emissions total $14-50 billion; in comparison, these LPG subsidies could be expected to cost $0.6-4.8 billion (Table 1, Fig 1d).

## Encouraging clean cooking through the removal of a value added tax in Kenya

In Kenya, LPG use has grown in the last fifteen years: from 4% of households primarily using LPG in 2006, to 13% in 2016, to 30% in 2022. In recent years, Kenya has experimented with a value-added tax on LPG: first, the long-standing 16% VAT was reduced to 0% in 2016 to encourage LPG adoption; it was then re-established at 16% in 2021 in response to the financial pressures of COVID-19; most recently, in July 2022, the VAT was halved to 8% to enhance affordability during international petroleum price surges.

To model the health and climate benefits of the potential removal of the VAT between 2023-2030, we first generate estimates of yearly LPG consumption among LPG users in Kenya. We then map LPG consumption to mean personal PM_2.5_ exposures, drawing on Kenya-specific estimates (Fig 1e). Next, we use observational evidence of within-household declines in LPG consumption due to the reinstatement of the VAT in 2021 to infer household price sensitivities. We apply these price sensitivities (1) to a case where the 16% VAT is removed and prices decline by proportionally and (2) to an alternative scenario where the VAT is 8%. Given that historical evidence suggests that the removal of the VAT may encourage further LPG adoption among current non-adopters, we additionally simulate varying levels of increasing LPG adoption at 0.5% (baseline), 1% (8% VAT), and 1.5% (0% VAT) per year. We then relate population shifts in LPG consumption to changes in pollution exposures and to relative risks of mortality, and then apply these estimates to predicted yearly crude mortality rates and population data to estimate changes in mortality. We similarly infer changes in biomass consumption and estimate changes in CO_2_e emitted due to the VAT removal using energy equivalences.

In the absence of the VAT, 30,000 premature deaths would be averted between 2023-2030 in Kenya (Table 1), decreasing national household air pollution related mortality by 20%. Net CO2e would be reduced by 7 megatonnes; biomass use is less renewable in Kenya, so it is relatively more emitting than other case studies due to lower rates of CO_2_ re-absorption.

Averted mortalities are equivalent to $9.8 billion and averted CO_2_e to $7.8 billion (Fig 1f). If the VAT were halved, our estimates are reduced by one-third. Still, the VAT is an economic tool to generate capital; based on the amount of predicted LPG consumption, we estimate that the 16% VAT would be expected to generate $740 million in revenue between 2023-2030 (~0.1% of expected government spending) (Fig 1f).

## Conclusions

Some caveats apply to our analysis. Our averted mortality estimates are sensitive to our choice of exposure-mortality relationship ^17^, our ability to estimate exposures under different cooking fuel scenarios, and on population and mortality data. Monetized emissions and subsidy costs are also subject to error owing to data constraints. We focus on readily quantifiable benefits (mortality and emissions), and do not account for other benefits of cleaner cooking including women’s empowerment, reduced healthcare expenditures, and to local environments^18^. We quantify costs related to directly subsidizing fuels, but broader investments along the fuel supply chain may be necessary to support growth in LPG use, though these may also offer opportunity for local job growth. Given uncertainties associated with our assumptions – which we aim to quantify through bootstrapping – we focus on the direction and magnitude of relative differences between costs and benefits as opposed to individual values.

Given the popularity of gas subsidies among biomass users, it is perhaps unsurprising that their removal – typically motivated by budgets – can be challenging. Previous efforts to remove gas subsidies in the three countries described here have been unpopular; as such, governments may understandably be reticent to consider such subsidies. Further, those who oppose fossil fuel subsidies argue that their removal is pro-climate, pro-health, and pro-poor^19,20^; for cooking gas subsidies, we contend otherwise.

While gas is usually better for climate and health than biomass, electricity powered by renewable sources is a better option still. Thus, some argue for a transition directly from biomass to electricity. We do not view subsidizing gas and encouraging electric cooking as mutually exclusive paths. Many around the world, in both developing and industrialized nations, use gas and electricity for cooking, e.g., they have a gas stove, a microwave, and a hot water kettle. Further, the supply chains for electricity and gas are complementary. As observed during the Covid-19 pandemic, gas cylinder delivery can be interrupted when vehicle movement is restricted, which can lead to increased reliance on biomass ^21^; electricity networks are resilient to these restrictions. At the same time, in many regions where biomass remains prevalent, electricity provision may at times fail or be insufficient, in which case ‘falling back’ to gas is a better option than reverting to biomass.

Gas, a transitional fuel available at scale now, is an intermediate step toward the better solution of cooking with electricity from clean, renewable sources. For those where that ideal is already an option, it should be aggressively pursued. Unfortunately, for many around the globe, that ideal is decades away. Until then, targeted and subsidized fossil fuels can fulfill the promise of healthier lives.

## Methods

### Estimating changes in mortalities

Our modeled estimates of the averted mortality from clean cooking fuel scale up in Ecuador rely on the fraction of households primarily using a clean cooking fuel linearly interpolated between decennial census years, predicted primary clean cooking fuel use absent the subsidy which approximates the observed data lagged by 20 years, all-cause all-age mortality rates from the World Health Organization, average PM_2.5_ exposure estimates for those using clean cooking fuels primarily and those that are not based on in-country personal air pollution exposure monitoring drawn from truncated normal distribution where means were 50 *μg/m*^3^ (sd = 20 *μg/m*^3^) for primary polluting fuels and 25 *μg/m*^3^ (10 *μg/m*^3^) clean (see Fig 1), and an exposure-response function that translates those exposures into changes in all-age all-cause mortality risk – the Global Exposure Mortality Model (GEMM) (see Supplement).

For India and Kenya, we rely on the logic that increases in LPG cylinder refill prices will reduce LPG consumption and increase biomass combustion. When biomass combustion increases, personal PM2.5 exposures increase, health risks increase, and all-cause, all-age mortality increase. Similarly, CO2e emissions go up because biomass stoves are less efficient and emit greenhouse gases more than LPG per unit energy delivered. To estimate mortality changes for plausible LPG cylinder refill price changes, we draw on distributions of LPG consumption from household surveys, an empirically-derived LPG price elasticities from (quasi-)experimental in-country studies^16,22^ (we assume that higher-consuming households are wealthier and more price inelastic), in-country personal PM_2.5_ exposure modeling among households using levels of LPG use (see Fig 1), population and crude mortality rate projections from 2023—2030, and the GEMM exposure-response relationship (see SM).

The Value of a Statistical Life is a dollar value that is meant to represent the aggregated, population-level willingness to pay for reductions in mortality risks – it is typically scaled to local contexts. While the US Environmental Protection Agency suggests that when conducting cost-benefit analyses one uses a central estimate of $7.4 million ($2006), updated to the year of analysis, we identify country-specific estimates for VSLs. In Ecuador, we select a preferred VSL of 820,000 USD ^23^, in India we select 640,000^24^, and Kenya 230,000^24^. We apply social discount rates of 5% throughout, and test alternative VSLs (see Supplement).

We bootstrap each case study 1,000 times drawing from distributions for LPG use – exposure relationships and also, for India and Kenya, in the price elasticities; values reported represent the mean.

### Changes in carbon emissions

We estimate total energy consumption from each fuel and then translate these combustion estimates to emissions using standard assumptions about daily energy consumption, fuel-specific combustion emissions, and the fraction of biomass that is renewably harvested (fNRB) using a reduced form of the approach outlined in Floess et al. (2023) ^10^ (see Supplement).

Kilograms of biomass are estimated as a direct function of LPG via energy equivalences and stove efficiency. In each case study, we estimate 1,000 iterations of kilograms of LPG and biomass combusted in each year in both scenarios, translate these to CO2e emitted, and take the difference across counterfactuals. We rely on Burke et al. (2023)^25^ to monetize changes in CO2 emissions in all three case studies. Year-specific values range from $379 / tCO2 in 1980 to $203 / tCO2 in 2020; future damages are discounted as described in the mortality modeling section.

## Data Availability

All data produced in the present study are available upon reasonable request to the authors and will be made publicly available upon publication.

## Acknowledgments

We gratefully acknowledge inspiration from the work of and conversations with Kirk R. Smith, Gautam Yadama, and Josh Rosenthal, among others.

## Data and code availability

Data and code to replicate all results in the paper will be made available upon publication.

## Funding

US National Institutes of Health grant 1UM1HL134590 in collaboration with the Bill and Melinda Gates Foundation (OPP1131279) (AP)

Clean Cooking Implementation Science Network of the US National Institutes of Health (AP)

## Author contributions

Conceptualization: CFG, AP

Methodology: CFG, AP, SBS, MB, RB

Formal Analysis: CFG, AP, SBS

Resources: SBS, RB

Writing – Original Draft: CFG, AP, SBS

Writing – Review & Editing: All

## Competing interests

Authors declare that they have no competing interests.

## Notes

### Competing Interest Statement

The authors have declared no competing interest.

### Funding Statement

US National Institutes of Health grant 1UM1HL134590 5 in collaboration with the Bill and
Melinda Gates Foundation (OPP1131279) (AP)
Clean Cooking Implementation Science Network of the US National Institutes of Health
(AP)

### Summary of Updates

Improved and clarified analysis approach (in particular for India and Kenya scenarios), added Figure 1.

