## Supplemental Information for "In praise of fossil fuel subsidies (for cooking) ^‡^"

### Context of cooking fuel programs in Ecuador, India, and Kenya

It is worth directly comparing the context of LPG and related programs in Ecuador, India, and Kenya. In Ecuador, LPG cylinder refill costs for all residential consumers are directly subsidized at point of sale (subsidy is ~$0.67/kg, 90% market cost). Since there is no restriction on the number of cylinder refills that can be purchased for most of the country, this universal and untracked subsidy leads to disproportionate subsidy capture by the wealthy (though use among the comparatively less wealthy is still high). There is also some leakage, both to neighboring countries and non-residential users. In India, LPG cylinder refills are purchased at market rates and subsidies are subsequently deposited into customers’ bank accounts (~$0.33/kg, 45% market cost). LPG cylinder refill costs are centrally dictated each month and there is little variation in costs within states. Eligible (income-poor) customers can avail themselves of nine subsidized refills yearly. India’s direct-deposit approach limits leakage and can enable flexible subsidy targeting; however, it also introduces barriers for the unbanked and for those that struggle to pay the upfront cost. In Kenya, LPG prices are not controlled and are directly influenced by international markets. As part of efforts to expand LPG use to 35% of the population primarily cooking with LPG by 2030 (from 20% in 2016), the Kenyan Treasury removed the 16% VAT on LPG, which effectively served as a 16% price cut for consumers. However, the 16% VAT on LPG was reinstated in 2021 amid COVID-19 related budgetary strain. As a result, LPG prices rose to $1.62/kg and have recently reached $1.73/kg.

### Ecuador case study

#### Data sources

Here we list some parameters and data sources in more detail than in the main text.

- Ecuador’s historical LPG sales volume and subsidies data come from multi-annual reports by PetroEcuador, the state petroleum company which, together with its predecessor entities, has held a monopoly on LPG importation and wholesaling throughout the period under study. Consumer prices can be found on [p.170](https://www.eppetroecuador.ec/wp-content/uploads/downloads/2015/03/40-A%C3%B1os-Construyendo-el-Desarrollo-del-Pa%C3%ADs.pdf) and annual consumption can be found on [p.102](https://www.eppetroecuador.ec/wp-content/uploads/downloads/2015/03/40-A%C3%B1os-Construyendo-el-Desarrollo-del-Pa%C3%ADs.pdf) of PetroEcuador report ‘40 years building the development of the country, 1972–2012.’
- Exchange rates between the US dollar and Sucre (Ecuador’s national currency until early 2000, when the country began using the US dollar as its official currency) were calculated using data downloaded from the Banco Central de Ecuador [(here)](https://contenido.bce.fin.ec/documentos/MercadosInternacionales/Cotizaciones/tipoCambio.xls). Conversion factors were applied to convert Sucres into current-year US dollars, and to convert current-year dollars into May 2023 dollars (Bureau of Labor Statistics [here)](https://www.bls.gov/data/inflation_calculator.htm).
- Ecuador’s historical GDP data come from the World Bank (available [here)](https://data.worldbank.org/indicator/NY.GDP.MKTP.CD).
- The Central Bank also collects and publishes data from PetroEcuador, including quarterly statistics on annual LPG imports and sales, which were used to crosscheck and fill gaps in the data from PetroEcuador-published reports.
- The cost of Ecuador’s LPG subsidy was determined by multiplying the country’s annual consumption by the difference between the subsidized consumer price and the international wholesaler and producer price (data from [EIA](https://www.eia.gov/dnav/pet/hist/LeafHandler.ashx?n=pet&s=ema_epllpa_pwg_nus_dpg&f=a) and [FRED](https://fred.stlouisfed.org/graph/?id=APROPANEMBTX) producer price index; propane using Dec 2022 as index=100).

This analysis makes several assumptions to bridge data gaps: first, domestic LPG production is treated as equivalent to imported fuel. Domestic production volume has been stagnant since the mid-1990s and has accounted for roughly one-fifth of overall consumption since 2000. As domestic production has largely come under the aegis of PetroEcuador, we treat unrealized income (from the potential sale of this LPG at market prices) as equivalent to the cost of imported fuel. Second, our estimations conservatively treat all LPG sales within the country as fully subsidized. While household use of LPG represents most of the internal market (>90% over the last decade), a complex and evolving set of price points have been fixed for industrial, agricultural, and transportation uses of LPG. However, data gaps on LPG use in these sectors for the majority of the years under analysis prevent the inclusion of this detail. Finally, our use of average annual international wholesale prices and US prices received by LPG producers for the period prior to 2011 represents an imperfect estimate, and likely an underestimate, of the cost paid by Ecuador’s government for imported LPG, which must cover shipping and logistics costs, in addition to being subject to the volatility of the LPG market at the moment that import contracts are signed.

#### Modeling mortality and estimating its value, 1979-2019

Our modeled estimates of the averted mortality from clean cooking fuel scale up in Ecuador rely on yearly nationwide observed % primary clean cooking fuel use, predicted % primary clean cooking fuel use absent the subsidy, all-cause all-age mortality rates, average PM_2.5_ exposure estimates for those using clean cooking fuels primarily and those that are not, and an exposure-response function that translates those exposures into changes in all-age all-cause mortality risk.

The observed trajectory is derived by linearly interpolating the percent primary clean cooking fuel use variable derived from Census years 1974, 1982, 1990, 2000, 2010, and a combined set of nationally-representative surveys conducted between 2015– 2019 (whose mean value we place at 2019). The 20 year delayed trajectory is calculated by starting at 6.55% in 1974 (no difference to observed), increasing to 9.91% in 1984 (the value Ecuador had reached in 1975), increasing to 26.71% by 1994 (1980), increasing to 39.57% in 2004 (1984), and maintaining the 20y delayed gap thereon. The 10y delayed trajectory is calculated by starting at 6.55% in 1974 (no difference to observed), increasing to 23.35% in 1984 (the value Ecuador had reached in 1979), increasing to 39.57% by 1994 (the value reached in 1984), and maintaining the 10y delayed gap thereon.

Yearly nationwide data on population and mortality are derived from the United Nations World Population Prospects 2022 (available [here)](https://population.un.org/wpp/Graphs/DemographicProfiles/Line/218).

Exposure contrasts are derived from ref. (*2*) and are as follows: 50 *µg/m*^3^ (sd = 20 *µg/m*^3^) polluting and 25 *µg/m*^3^ (10 *µg/m*^3^) clean. For each bootstrapped run, we draw from a truncated normal distribution where the mean and SD are as described above, the minimum is 5 *µgm*^3^ for both polluting and clean fuels, and the maximum is 150 *µgm*^3^ for polluting and for clean fuels. The maximum was the randomly selected level for the polluting group, ensuring that the clean group had either the same exposure or lower.

For the exposure-response relationship (for Ecuador, India, and Kenya), we applied the Global Exposure Mortality Model (GEMM).(*3*) The GEMM provides hazard functions derived from 41 cohort studies across 16 countries, including high PM2.5 contexts, and removes the use of secondhand smoking studies to cover these high concentrations and to provide an upper exposure bound. Our outcome of interest is nonaccidental mortality; we apply the 25 years and older category from the GEMM to our populations. The inputs are drawn from an excel file downloaded from the Supplemental Information from the paper describing the GEMM and its application (Burnett et al. (2018)).

As noted elsewhere,(*4*, *5*) the choice of exposure-response relationship can highly impact modeled estimates of air pollution attributable mortality, especially at high concentrations (by 2x, at times). In addition to its strengths noted above, we choose the GEMM because its parameters are publicly available and easily implemented.

We select a preferred Value of a Statistical Life (VSL) of 820,000 USD from (*6*). Other values are possible, however: 400,000 USD in 2019 dollars (*7*) and 2.15 million USD in 2020 dollars (*8*). For reference, in the US, The Department of Health and Human Services recommends using an $11.4 million VSL, The Department of Transportation recommends an $11.7 million VSL, and the Environmental Protection Agency recommends a $10.8 million VSL (US$2020).

#### Regression-based estimates of health benefits, 1990–2019

As a complement to our theoretical analysis detailed above, we additionally conducted regression-based estimates of health benefits from 1990 to 2019. This analysis builds on previous work^1^ where we estimated the impact of LPG scale-up from 1990 to 2019 on under-5 lower respiratory infections mortality (but is unpublished elsewhere). In comparison to Gould et al. (2023)(*9*), our approach differs in a few respects. First, our outcome of interest is all-cause, all-age mortality to more fully capture the potential benefits of clean fuel scale up. Second, to quantify uncertainties, we bootstrap 95% confidence intervals by sampling cantons with replacement. Third, we generate population averaged estimates by weighting canton-months according to their population in our regression.

To model the relationship between all-cause mortality and clean fuel, we estimate the following regression:

*log*(*ycy*) = *βPcy* +*λcy* +*µc* +*γy* +*θcy* +*εcy*	(1)

using ordinary least squares, where *c* indexes cantons and *y* indexes year. *y_cy_* is the log of the yearly canton-level all-cause, all-age mortality rate and *P_cm_* is the proportion of households primarily cooking with a clean-burning cooking fuel in the same canton-year. *λ_cy_* is a vector of canton-year control variables, including the fraction of households that are classified as rural, the fraction of households that are grid electrified, a composite index of household building materials (roof, wall, floor) intended to serve as a proxy for infrastructure and wealth, toilet type intended to serve as a proxy for water, sanitation, and hygiene practices and wealth, the fraction of adult women that are literate, the fraction of girls under 18 years that are in school, the fraction of households where an individual speaks an indigenous language, and a composite index of under-5 vaccination rates (for more details see ref. (*9*)). *µ_c_* is a vector of canton fixed effects that account for all locality-specific time-invariant characteristics correlated with either LPG scale up or mortality rates. To account for longer-term trends in LPG scale up and mortality rates, we include a vector of year fixed effects *γ_m_*, which account for any time-trending differences in either LPG scale up or mortality rates that are common to all cantons. Regressions were weighted by canton population and standard errors were clustered at the canton level.  We find that all-cause mortality rates decline by 0.41% (95% CI, -0.94% to 0.00%) with each additional percentage of households in a canton using a clean cooking fuel.

One might also be concerned that our results are sensitive to our selection of control variables. While these are theoretically motivated to cover multiple domains that might be correlated with both all-cause mortality rates and clean cooking fuel scale-up from 1990–2019 and the canton-year level, there are some others that could be selected as well. We test the full range of combinations of our included control variables as well three more (the fraction of pregnant women that received antenatal care, the fraction of children under 5 years that received three doses of the pneumococcal conjugate vaccine (introduced in 2010), and a composite index of household sanitation practices (source of household water, trash disposal practices, toilet type, and presence of a private household shower). In total, there were 2,054 combinations. Per 10 percentage point increase in the fraction of households primarily cooking with a clean burning cooking fuel, we observed a decline in all-cause mortality rate of an average of -5.4% (the median was -4.9% (IQR, -2.7% to -7.6%)) across all combinations of controls.

#### Benchmarking estimates with the Global Burden of Disease

We benchmark our two approaches using estimates from the Global Burden of Disease (GBD). The GBD estimates mortality from household air pollution by collecting data on exposure, health endpoints, and population demographics and applying these country-level statistics to modeled concentration-response functions over time. GBD models indicate that 27,000 more deaths would have occurred had household air pollution levels stayed fixed from 1990–2019, i.e., 30 years of 1990 household air pollution mortality estimates exceed the ‘observed’ household air pollution related mortality estimates by 27,000 deaths. Our first approach based on vital statistics yields an estimate of 34,500 excess deaths; our second model-based approach estimates 39,000 excess deaths.

### India case study

The central reference provided in the main text is to India’s energy budget for 2023-2024. In conjunction with budgets since 2016-2017, it is clear that investments in PMUY and subsidies have declined precipitously (see [collated budget data)](https://scroll.in/article/1043498/modis-lpg-subsidy-programme-is-not-as-big-a-success-as-the-finance-minister-suggests). On August 29, 2023, the Government of India announced a cut to domestic LPG cylinder refill prices of roughly 200 INR, in response to rising inflation and high international petroleum prices; this move also comes amid ramp up for 2024 national elections in India. This brings the total subsidy rate for PMUY-eligible households to 400 INR per cylinder; other consumers receive 200 INR per cylinder. Up-to-date LPG cylinder refill prices can typically be found at <https://www.goodreturns.in/lpg-price.html>.

Central to our estimates of the health-related impacts of cooking gas subsidies in India is the logic that if the LPG cylinder refill prices increase, then LPG cylinder refill purchasing (and thereby consumption) decreases. When LPG consumption decreases, biomass combustion increases. When biomass combustion increases, personal PM2.5 exposures increase, health risks increase, and at the population level public health is damaged. Similarly, given that biomass emits more greenhouse gases than LPG per unit energy delivered, CO2e emissions increase. Similar logic applies to the reverse, which we model directly: lower prices lead to more LPG consumption and more biomass displacement. Here, we outline specific inputs for our modeling and our approach.

*LPG prices*

In 2019, LPG cylinder refills cost 750 INR, subsidized to 550 INR for PMUY beneficiaries. As of September 2023, LPG cylinder refills cost 900 INR, subsidized to 700 INR for PMUY beneficiaries. However, absent the recent price cut, costs would be at 1100 INR, subsidized to 900 INR for PMUY beneficiaries. Previously, budgets indicated that there would be minimal support for the LPG subsidy, indicating that it would be plausible that PMUY beneficiaries would have to pay 1100 INR per refill. As such, we model three scenarios – 1100 INR, 900 INR, and 700 INR -- and compare them to 1100 INR costs.

*LPG price elasticity*

To estimate the extent to which LPG consumption declines with increases in refill prices, we draw on recent experimental work in Tamil Nadu, India among PMUY beneficiaries (*10*). Briefly, households recruited into the study were randomized to receive differing levels of subsidies for LPG cylinder refills and their refill and LPG consumption were tracked over time. Results from that study indicate strong price sensitivity, which is supported by rich observational evidence from both India and elsewhere globally. We estimate price sensitivities using results presented in Table 2 of that study, which show the mass of LPG consumed during the intervention period in kilograms across the control group and various subsidy levels ($1.70, $3.40, $5.10). Using a baseline control cost of about $7.95 per refill, we estimate price elasticity among PMUY beneficiaries by regressing the log of consumption per month (estimated as group averages divided by an intervention period length of seven months) and the log of the price. This procedure yields an estimated price elasticity of -0.33. In other words, a 1% increase in LPG cylinder refill costs results in a decline in LPG consumption of 0.33%.

Given that high consuming households also have larger incomes, we infer that they are less sensitive to price than low-consuming households—for whom a single LPG cylinder refill accounts for a larger fraction of their monthly expenditures. As such, in our preferred specification, households that consume fewer than 4 refills per year have an elasticity of -0.33, 4-9 refills per year have an elasticity of -0.2, 9-12 refills per year have an elasticity of -0.1, and those than consume more than 12 refills per year are price inelastic. In Scenario 2, all households have a price elasticity of -0.33. In scenario 3, all households have a price elasticity of -0.10. In analyses, we randomly draw elasticity from relatively tight means/SDs for all; each elasticity must be a lower step down and never 0 (except for those than consume >12 refills per year). When reporting results, we refer to these three scenarios as “Trade-off Scenario 1”, “Trade-off Scenario 2”, and “Trade-off Scenario 3.” LPG price elasticities are referred to as Elasticity 1, 2, or 3 (in our main model they are -0.33, -0.2, and -0.1, respectively). Random draws are specified as follows (R code; rnormTrunc comes from the *EnvStats* package):

elasticity1 = rnorm(1, .33, .025)

elasticity2 = rnormTrunc(1, .2, .025, 0.1, elasticity1)

elasticity3 = rnormTrunc(1, .10, .0025, 0.01, elasticity2)

LPG consumption

LPG consumption is derived from energy access survey data collected in 14,850 urban and rural households across 152 districts in India’s 21 most populous states in 2019 (*11*). A stratified multistage probability sampling design was implemented to achieve nationally representative data when accounting for household level survey weights. All households were asked if they had an LPG stove. If so, they were asked if they obtained their stove via PMUY. Households that had an LPG stove were also asked how many 14.2 kg LPG cylinder refills they purchased in the previous year. As such, we can estimate the distribution of LPG consumption in kg per year among PMUY beneficiaries.

We compute LPG consumption in a few steps. First, we (1) generate a 2019 density plot; (2) extract values at every 1kg between 1-200 kg/year; and (3) scale densities to sum to 1 so that every value at a 1kg increment can be considered as a % of population. Next, we estimate our ‘baseline LPG consumption’ by, for each 1kg/year, multiplying the % price change (relative to 550 INR) by the price elasticity to get declines in LPG consumption. In this ‘baseline’ scenario, LPG cylinder refill prices are 1100 INR.

To estimate the benefits of the LPG subsidy, we then re-model LPG consumption as a function of anticipated LPG cylinder refill price declines (and the price elasticities) relative to 1100 INR. For each of these scenarios (where prices are 550 INR, 700 INR, and 900 INR), we subtract kg declines from the estimated consumption in the 2023 kg/year distributions.

*PM_2.5_ exposures*

There are no recent, nationally representative Indian personal air pollution exposure measurements for households that use exclusively polluting cooking fuels, that mix polluting and clean-burning fuels, and that use exclusively clean-burning fuels. However, a recent and extensive set of measurements in Tamil Nadu, India – of pregnant women, children, and other adult women in the same households – provides evidence of the effectiveness of LPG in reducing exposures when used consistently. In refs. (*12*, *13*) biomass using households had mean PM_2.5_ exposures of 100-120 *µg/m*^3^, while LPG using households had median exposures ranging between 37-39 *µg/m*^3^. We note that the exposures observed in Tamil Nadu are lower than the expected range for biomass using households across India, but nonetheless utilize these estimates as they result in conservative overall modeled averted mortality estimates.

We develop three scenarios to relate LPG consumption to average PM2.5 exposures. For each of these models we develop a simple formula with parameters that we randomly draw from in each bootstrapped run; the LPG to PM2.5 exposure relationship varies slightly across all runs. In each run, we model all three LPG-PM2.5 relationships fully. The central responses include:

(a) Our preferred, sinusoidal response where there is little movement in exposure until about two to three refills per year, then a steep decline until about eight or nine, at which point the response levels off.

(b) A sinusoidal response that can be considered to be somewhat more pessimistic about how much LPG consumption results in declines in exposure. In other words, more LPG consumption is needed to effectively reduce PM2.5 exposures.

(c) A linear response from the maximum exposure (1 kg) to the minimum (200 kg).

In each bootstrap we run all three scenarios and perturb their parameters somewhat (slightly higher/lower max/min, earlier/later, steeper/shallower declines). Specific R code to do so is shown below:

min1 = rnorm(1, 25, 5)

max1 = rnorm(1, 100, 5)

decline1 = rnorm(1, 70, 2.5)

exp1 = rnorm(1, 4, .05)

exp2 = rnorm(1, 8, .1)

x=0:200

PM25_a=min1 + (max1-min1) / (1 + (x/decline1)^exp1)

PM25_b=min1 + (max1-min1) / (1 + (x/max1)^exp1)

PM25_c=max1 - ((max1 - min1) / 200)*x

*Other model inputs and parameters*

We use predicted data on the size of India’s population and crude mortality rate yearly from 2023–2030. These data are derived from the United Nations World Population Prospects 2022 (available [here)](https://population.un.org/wpp/Graphs/DemographicProfiles/Line/218). We note that the UN WPP indicates that India’s crude mortality rate will decline from 9.1 per 1000 in 2022 to 6.6 per 1000 in 2023. If the crude mortality rate does not drop to this extent, our results may underestimate mortality damages due to higher-than-expected baseline mortality rates, though ex ante it is difficult to know how this may alter the effect of LPG subsidies on the margin.

Our choice of *VSL* (820,000 USD) comes from ref.^13^

Given that we are projecting future benefits, it is useful to apply a social discount rate. According to ref.(*14*) based on growth rates, a discount rate of 9% is appropriate for India in global health analyses.

Our choice of time horizon (2023–2030) is aligned with the Sustainable Development Goals ‘deadline’ of 2030, though, of course, is somewhat arbitrary. We refrain from predicting beyond 2030 to avoid even larger uncertainties in fuel markets and other policies and political environments.

#### Mortality modeling

To estimate mortality damages, we estimate the number of people at each 1 kg/year increment, which is directly mapped to personal PM2.5 exposure, as outlined above. For a given level of PM2.5 exposure, we use the GEMM to identify the mortality hazard ratio for that kg/year population group. Applying this hazard ratio to the national crude mortality rate, we then estimate excess mortality relative to ‘theoretical minimum’ 2.4 ug/m3. We then sum yearly excess mortality due to PM2.5 exposure within a year and estimate differences across scenarios within a year. We then apply VSLs (preferred, high, low), apply the social discount factor, and summarize across various model parameters: price, price sensitivities, and LPG consumption to PM2.5 exposure mapping.

### Kenya case study

Our approach to modeling health and climate benefits changes in VAT mirrors that of the India analysis.

#### Data sources

*LPG consumption*

Recent estimates of LPG ownership and use are derived using two data sources. First, the use of LPG as a primary cooking fuel comes from the Demographic Health Survey conducted in 2019, which is nationally-representative. One limitation of this survey is that we lack robust data on the use of secondary cooking fuels (either LPG or biomass). We complement these DHS data with data from Shupler et al. (2021)(*15*) who conducted 1840 household energy use surveys in Eldoret, Kenya. More specifically, we use data from 757 randomly-sampled households in Eldoret. These data indicate that among LPG users, 4% of households are exclusive LPG users, 37% are primary LPG / secondary biomass, and 59% are primary biomass / secondary LPG. In total, we estimate that 31.2% of households in Kenya use LPG in some capacity.

Shupler et al. (2021) also report kilograms of LPG consumed per capita per year across the different LPG use types. We simulate normal distributions of LPG kg/capita/year across the different household samples to generate hypothetical populations to recover the ‘raw’ data from this study. By combining these distributions with data on the number of households in Kenya, we can recover distributions of LPG consumption (kg/capita/year) for all Kenyan households.

*LPG prices*

We model three central scenarios: no VAT, VAT of 8%, and VAT of 16% (reference). We estimate percent change based on the removal (or reduction) of the VAT relative to current prices. Based on a reference price of $1.73/kg, which internalizes the 16% VAT, the price reduction in percentage terms would be 14%. For the 8% VAT scenario, the price reduction is 7%. This agrees with observational evidence from Shupler et al. (2023) (*16*).

*LPG price elasticity*

To estimate the extent to which LPG consumption declines with increases in refill prices, we draw on recent work by Shupler et al. (2023), who observed LPG consumption records among a subset of households that use pay-as-you go technologies. Using real-time data on LPG prices and consumption before and after the VAT was re-established, they estimate a change in LPG consumption with a change in price. Specifically, Shupler et al. (2023) report^[2]^ that LPG prices increased by 34 Kenyan Shillings (KSh) from 214 to 249 KSh per kilogram, which was accompanied by a decline in LPG consumption of 0.55 kg/capita/month (from 0.82 kg/capita/month). We thus estimate that a price increase of 16% resulted in an LPG consumption decline of 67%. Using these percent changes, we recover a price elasticity of 4.1. Elsewhere, Shupler and colleagues report that wealthier households (and higher consuming households) were less likely to alter LPG consumption as a result of the VAT changes (*17*). We model price elasticity differently across consumption groups, where those that are highest consumers have the lowest elasticity and vice-versa. We again draw randomly from a distribution in each bootstrap run. R code is as follows:

elastic3 = 4.101045/10 # >30 kg/capita/yr

elastic2 = 4.101045# 15-30 kg/capita/yr

elastic1 = 4.101045*1.1 # <15 kg/capita/yr

elasticity1 = rnorm(1, elastic 1, .025)

elasticity2 = rnormTrunc(1, elastic2, .5, 0.1, elasticity1)

elasticity3 = rnormTrunc(1, elastic3, .0025, 0.01, elasticity2)

*PM_2.5_ exposures*

Recent, nationally representative personal air pollution exposure measurements for households that use exclusively polluting cooking fuels, that mix polluting and clean-burning fuels, and that use exclusively clean-burning fuels in Kenya are not available. Instead, we draw on a set of other studies to establish baselines for these categories.

Drawing on the PURE cohort study, Shupler et al. (2020)(*18*) establish average PM2.5 exposures (medians and 25^th^-75^th^ percentiles) for households in Eastern Sub-Saharan Africa as follows. They estimate relatively high levels of exposure (in µgm^-3^): among gas/electric users, median female, male, and child exposures (25^th^, 75^th^ percentiles) are 102 (34, 314), 73 (24, 226), and 89 (30, 273), respectively, indicating that clean fuel uses in these categories may not be exclusively using these fuels. For wood users, median female, male, and child exposures (25^th^, 75^th^ percentiles) are 388 (115, 1122), 279 (83, 808), and 337 (100, 976), respectively.

Elsewhere, a study in Kenya focused on predicting personal exposures (Johnson et al. (2021)(*19*)) reports lower estimated predicted and measured exposures among LPG users and wood users (in µg/m3): median (25^th^, 75^th^ percentile) exposures of 29 (27-46); for wood users, they measured 182 (104, 292).

Using these inputs, we assign average PM2.5 exposure for exclusive LPG use to be roughly 35 µgm^-3^. Average PM2.5 exposure for exclusive biomass use is 250 µgm^-3^. Both of these estimates are perhaps optimistic and ultimately may mean we underestimate impacts. If we halve the exclusive biomass value in our models to 125 ug/m3, modeled mortality impacts are attenuated by 10-15%.

As with the India model, we develop three scenarios to relate LPG consumption to average PM2.5 exposures. For each of these models we develop a simple formula with parameters that we randomly draw from in each bootstrapped run, so the LPG to PM2.5 exposure relationship varies slightly across all runs. In each run, we model all three LPG-PM2.5 relationships fully. The central responses include:

(a) Our preferred, sinusoidal response.

(b) A somewhat more pessimistic sinusoidal response.

(c) A linear response from the maximum exposure (1 kg) to the minimum (55 kg).

In each bootstrap we run all three scenarios and perturb their parameters somewhat (slightly higher/lower max/min, earlier/later, steeper/shallower declines). Specific R code to do so is shown below:

```
min1 = rnorm(1, 35, 5)
max1 = rnorm(1, 250, 25)
decline1 = rnorm(1, 18, 2.5)
decline2 = rnorm(1, 13, 2.5)
exp1 = rnorm(1, 3, .05)
exp2 = rnorm(1, 5, .1)

x=1:55
PM25_1=min1 + (max1-min1) / (1 + (x/decline1)^exp1)
PM25_2=min1 + (max1-min1) / (1 + (x/decline2)^exp1)
PM25_3=max1 - ((max1 - min1) / max(x))*x

kg_lpg_pm25 <- tibble(
  kg_lpg = 1:55,
  PM25_1 = PM25_1,
  PM25_2 = PM25_2,
  PM25_3 = PM25_3
)
```

*Other model inputs and parameters*

We use predicted data on the size of Kenya’s population and crude mortality rate yearly from 2023–2030. These data are derived from the United Nations World Population Prospects 2022 (available [here)](https://population.un.org/wpp/Graphs/DemographicProfiles/Line/218).

Our choice of VSL (230,000 USD) comes from ref (*20*).

We apply a social discount rate; according to Haacker et al. (2019)(*14*) a discount rate of 5% is appropriate for low- and middle-income countries in global health analyses.

#### Mortality modeling

As in the India model, to estimate mortality damages, we estimate the number of people at each 1 kg/capita/year increment, which is directly mapped to personal PM2.5 exposure, as outlined above. For a given level of PM2.5 exposure, we use the GEMM to identify the mortality hazard ratio for that kg/capita/year increment population group. Applying this hazard ratio to the national crude mortality rate, we then estimate excess mortality relative to the ‘theoretical minimum’ 2.4 µgm^-3^. We then sum yearly excess mortality due to PM2.5 exposure within a year and estimate differences across scenarios within a year. We then apply VSLs (preferred, high, low), apply the social discount rate, and summarize across various model parameters: price change, price elasticities, and LPG consumption to PM2.5 exposure mapping.

### Estimating greenhouse gas emissions differences in all three case studies

To estimate the greenhouse gas emissions differences across scenarios we need to estimate total energy consumption from each fuel and then translate these combustion estimates to emissions. To do so, we follow common, simple assumptions about daily energy consumption, fuel-specific combustion emissions, and the fraction of biomass that is renewably harvested (fNRB) (and thus does not contribute to net emissions). Broadly we follow a reduced form of the approach outlined in Floess et al. (2023) (*21*).

We rely on a set of emissions factors that quantify upstream and at-point-of-combustion contributions of cooking fuels to CO2, CH4, and N2O) emissions, which are then converted to CO2-equivalent (CO2e) using global warming potentials (1, 27.8, and 273, respectively). We have these emissions factors for firewood and LPG in grams CO2e emitted per megajoule (MJ) delivered from fuel combustion. For firewood, we defray a portion of related emissions by multiplying it by the fNRB. Where feasible, we apply country-specific fNRB values (India = 27%, Kenya = 61.1%). Ecuador does not have a country-specific fNRB, so we use the low- and middle-income country average of 28.8%, which is similar to nearby countries’ values (Colombia 29.7%, Peru 26.1%).

#### Ecuador

In Ecuador, we model CO2e differences due to the subsidy on the basis of the fraction of the population primarily using LPG or biomass in a given year in the observed vs. 20-y delayed scenario. To do so, we estimate the total MJ delivered for each fuel in each household, then by the population size, and by the emissions factors to obtain CO2e estimates for each year.

*Biomass*

We estimate CO2e emissions for biomass in a given year using the following equation

*CO*2*e_biomass_* = *γ* ∗ *EF* ∗ *ω* ∗ 7.3 *MJ / person / day* ∗ 365 *days* ∗ *fNRB*	(2)

where *γ* is the total population using biomass and *ω* is the fraction of cooking that is done using biomass. The emissions factor (EF) is estimated from Floess et al. indicate that per megajoule energy delivered from biomass combustion the following are emitted: 3.87g CH4, 0g N2O, and 633.7g CO2. Given that these are per MJ delivered, they already take into consideration the energy efficiency of typical stoves. For LPG, this was 50%; for biomass it was 15%. Using global warming potentials (GWPs), we can estimate the equivalent emissions from CH4 and N2O to CO2 using standard factors of 28, 273, and 1, respectively. We estimate that per MJ delivered biomass combustion is associated with 741g CO2e. This then gets multiplied by the fNRB (28.8%), yielding estimates for Ecuador of 290g CO2e per MJ biomass delivered.

*LPG*

We estimate CO2e emissions for LPG each year using the following equation:

*CO*2*e_LPG_* = *γ* ∗ *EF* ∗ *ω* ∗ 7.3 *MJ / person/day* ∗ 365 *days*	(3)

where *γ* is the total population using LPG and *ω* is the fraction of cooking that is done using LPG. The emissions factor (EF) is estimated from Floess et al. (as with the biomass estimates) which indicate that per megajoule energy delivered from LPG combustion the following are emitted: 0.234g CH4, 0.0004g N2O, and 166g CO2. Following the above procedure, we estimate that, for Ecuador, LPG combustion is associated with 172.6 g CO2e per MJ delivered.

#### India

Our modeling procedure for estimating mortality impacts from increases in LPG cylinder refill prices can be used to generate estimates of the total kilograms of LPG consumed each year. When compared against the baseline scenario, this can be interpreted as changes in LPG kg consumed due to changes in LPG prices. We use these estimates of LPG kg changes to make inferences about greenhouse gas emissions. Using the specific heat of LPG of 45 MJ/kg, we can convert kg consumed to MJ. Using the above calculations, we can then easily estimate total kg CO2e from LPG combustion averted from reduced LPG consumption:

*CO*2*e_LPG_* = *γ* ∗ *45 MJ / kg* ∗ 0.173 kg CO2e / MJ

Where *γ* is additional kg LPG consumed per year.

While this implies reduced CO2e emissions due to LPG refill price increases, we must account for expected increases in traditional biomass combustion. Since reliable, empirical evidence of the substitution for LPG and biomass for cooking is in large part unavailable, our central approach to estimating concomitant increases in biomass combustion from reduced LPG consumption is through energy equivalences. First, we consider the efficiencies of traditional Indian chulha stoves (~15%) and LPG stoves (~51%). Dividing these two values can help us to estimate a trade-off if household demand for energy delivered were unchanged by shifting between biomass and gas (0.29). Thus, we multiply MJ of averted kg LPG by 0.29 to obtain an estimate of increased biomass combustion in MJ. For each additional MJ firewood combustion, we estimate 278.5 g CO2e/MJ firewood delivered.

#### Kenya

Our approach for modeling in Kenya is identical to the approach outlined for India, except that the fNRB in Kenya is substantially higher, which results in much higher GHG emissions from firewood cooking (529.9 g CO2e/MJ delivered).

#### Monetizing CO2 emissions

We rely on Burke et al. (2023)(*22*) to monetize changes in CO2 emissions in all three case studies. Burke et al. (2023) link recent efforts to quantify the harms of CO2 emissions through 2100 (i.e., the social cost of carbon; SCC), and generate year-specific SCCs from 1980 to 2022. These year-specific SCCs are a large advancement over previous efforts to quantify SCCs and are a central motivating factor for using these estimates. We use their 2% discount rate scenarios. In general, the SCC ranges from $379 / tCO2 in 1980 to $203 / tCO2 in 2020. For future damages, we discount the SCC by the social discount factor for each country as mentioned in the mortality modeling section.

6.	Sander,Klas,Mira-Salama,Daniel,Feuerbacher,Arndt, The cost of air pollution - a case study for the city of Cuenca, Ecuador. *World Bank*, (available at https://documents.worldbank.org/en/publication/documents-reports/documentdetail/458511468189273908/The-cost-of-air-pollution-a-case-study-for-the-city-of-Cuenca-Ecuador).

14.	M. Haacker, T. B. Hallett, R. Atun, *Health Policy and Planning*, czz127 (2019).

15.	M. Shupler *et al.*, *Nat Energy*. **6**, 1198–1210 (2021).

16.	M. Shupler *et al.*, “Declining use of clean cooking fuels & food security in 2022: Downstream impact of the Russian-Ukrainian war in a Kenyan informal urban settlement” (preprint, Epidemiology, 2023), , doi:10.1101/2023.07.09.23292423.

17.	M. Shupler *et al.*, *Applied Energy*. **292**, 116769 (2021).

18.	M. Shupler *et al.*, *The Lancet Planetary Health*. **4**, e451–e462 (2020).

19.	M. Johnson *et al.*, *Indoor Air*, in press, doi:10.1111/ina.12790.

20.	W. K. Viscusi, C. J. Masterman, *Journal of Benefit-Cost Analysis*. **8**, 226–250 (2017).

21.	E. Floess *et al.*, *Environ. Res. Lett.* **18**, 034010 (2023).

22.	M. Burke, M. Zahid, N. Diffenbaugh, S. Hsiang, “Quantifying Climate Change Loss and Damage Consistent with a Social Cost of Greenhouse Gases” (w31658, National Bureau of Economic Research, Cambridge, MA, 2023), p. w31658.

1. ^‡^ Our title draws inspiration from Kirk R. Smith (2002) ‘In praise of petroleum?’ *Science* and Kirk R. Smith (2014) ‘In praise of power’ *Science*.
2. Reported estimates come from personal correspondence from Dr. Matthew Shupler, who clarified that the cited preprint was out of date. Dr. Shupler provided these referenced numbers.
